# Evaluating the Efficacy of Tocilizumab in Moderate to Severe COVID-19 with Progressive Illness despite Steroids: Identifying the Optimal Timing of its Administration in C3G study

**DOI:** 10.1101/2020.11.07.20226837

**Authors:** Surabhi Madan, Manish Rana, Rohan Gajjar, Nitesh Shah, Vipul Thakkar, Bhagyesh Shah, Pradip Dabhi, Minesh Patel, Hardik Shah, Rashmi Chovatiya, Maulik Soni, Nirav Bapat, Amit Patel

## Abstract

**Background:** High mortality has been described in coronavirus disease 2019 (COVID-19) with cytokine release syndrome (CRS). Tocilizumab (TCZ), an interleukin-6 (IL-6) receptor antagonist may be associated with improved outcomes in such patients; however, the subgroups of patients who benefit the most need to be identified.

**Objective:** To analyze the efficacy and optimal timing of administration of TCZ in moderate to severe COVID-19 with features of CRS, where the response to steroids was poor.

**Methods:** This is a retrospective study of 125 patients admitted between May 5 to July 31, 2020, in a tertiary care hospital in western India, with moderate to severe COVID-19 who were treated with TCZ along with steroids. The primary outcomes were the need for mechanical ventilation (MV) or death, and secondary outcomes were a decrease in oxygen requirement and inflammatory markers; the incidence of secondary infections, and renal or hepatic dysfunction. Kaplan Meier survival analysis and log rank test were used for evaluating primary outcomes. Secondary outcomes were analyzed using the Wilcoxon Signed-Rank test.

**Results:** Among 1081 patients admitted during the study period, 125 were administered TCZ (median age, 56 [95% CI 54 - 60] years; 100 [80%] male). The commonest symptoms were fever (96%), cough (64%), and dyspnea (48.8%). 78.4% patients had comorbidities (hypertension 51.2%, diabetes 43.2%, obesity 25.6% and chronic cardiac disease 13.6%). Of 117 patients who were treated with TCZ before requiring MV, 18.8% progressed to MV. Overall, 25% of the patients needed MV support. 65.3% of patients were discharged by day 14 after TCZ administration. Mortality was nil, 16.2%, 50%, and 62.5% in patients who received TCZ on room air, low flow oxygen, high flow nasal cannula (HFNC) and bilevel positive airway pressure (BiPAP), and MV respectively; overall 24.8% of patients died. Survival analysis showed no difference in outcome with respect to age and gender, while progression to MV showed a statistically significant reduction for the event death (90.9% of patients who progressed to MV died as compared to 6.3% who did not; log rank test with p < 0.0001). No adverse events were noticed.

**Conclusion:** Mortality was least in patients of COVID-19 with CRS who received TCZ while on low flow oxygen. When administered in the early hypoxemic phase, TCZ is associated with reduced mortality and decreased need for mechanical ventilation.

## INTRODUCTION

COVID-19 is characterized by an initial phase of viral replication, followed, in a subset of patients, by a second phase of “hyperinflammatory syndrome” driven by the host inflammatory response to the virus.^1^ The most critical patients develop a so-called CRS or *cytokine storm*, with release of interferons, interleukins, tumor necrosis factors, and several other inflammatory mediators. This hyperinflammatory immune response is considered to be responsible for lung injury, and acute respiratory distress syndrome (ARDS), the major cause of mortality in COVID-19.^2^ High levels of IL-6 have been found to have a correlation with the severity and poor outcome of the disease.^3,4^ Also, IL-6 may be a significant contributor to thrombosis, having been associated with tissue and vascular endothelial cell injury, contributing to platelet aggregation and angiotensin II microvascular dysfunction.^5,6^ TCZ, a humanized monoclonal antibody that functions as an IL-6 receptor antagonist, has been used Off-label to mitigate the cytokine storm in patients with severe COVID-19. Various observational studies have provided insight into the role of TCZ in severe disease, where its use was associated with clinical improvement and decreased mortality. Though the results of the multinational, randomized placebo-controlled phase 3 trial evaluating TCZ in the treatment of severe COVID-19 pneumonia are underway (NCT04320615), the preliminary results of the French CORIMUNO-TOCI 1 trial suggest that a significantly lower proportion of the TCZ-group patients reached the primary composite outcome of need for ventilation (non-invasive or mechanical) or death at day 14.^7^ Similarly, the results of phase 3 EMPACTA trial showed that the patients who received TCZ were less likely to progress to mechanical ventilation or death.^8^

The objective of our study was to assess the course of illness and final outcome in a large cohort of patients with moderate to severe COVID-19 disease who received TCZ. Also, we aimed to analyze the optimal timing of administration of TCZ during the course of illness, which could be one of the key determining factors for the final outcome.

## METHODS

### Study design and Setting

This is a retrospective review and data analysis of 125 COVID-19 patients hospitalized in a 350 bedded, tertiary care private hospital in the western India; who received TCZ during the study period, from May 5 2020 to July 31 2020. The study was approved by the CIMS (Care Institute of Medical Sciences) hospital ethics committee (CTRI/2020/05/025247). The need for consent was waived off due to the nature of the study.

### Treatment protocol

We started admitting COVID-19 patients in the first week of May 2020. A CIMS COVID CARE GROUP (C3G) comprising of infectious diseases consultant, pulmonologists, intensivists, and physicians was formulated. Though there were minor inter-consultant variations, the C3G followed a uniform treatment protocol; based on the categorization of patients into mild, moderate, and severe categories, as defined by the Government of India, wherein mild disease is defined as the presence of upper respiratory infection without evidence of hypoxia, moderate disease is the presence of clinical features suggestive of pneumonia with SpO2 <94% (range 90-94%) on room air; and severe disease is clinical signs of pneumonia with SpO2 <90% on room air.^9^

The treatment protocol continued to be updated regularly, as per the availability of new information regarding various drugs, which mainly included hydroxychloroquine (HCQ), azithromycin, remdesivir, anticoagulants and statins. All the patients received standard of care treatment, either HCQ (alone or in combination with azithromycin) in the early part of pandemic or remdesivir (once it got available, after which the use of HCQ became almost nil). However, the indications and use of steroids and TCZ remained uniform throughout. TCZ availability was uninterrupted in our hospital.

The indications of treatment with TCZ were persistent or worsening hypoxemia (SpO2 <94% on room air); or persistent high-grade fever with increasing inflammatory markers, especially C-reactive protein (CRP) and ferritin; or IL-6 more than 5-7 times the upper normal limit (UNL) despite treatment with steroids for 48 hours. Almost all patients with moderate to severe disease received steroids, either methylprednisolone (MPS) or dexamethasone, before the initiation of TCZ, except those who presented to the hospital in cytokine storm with severe hypoxia, where TCZ was given on the same day as steroids. The dose of TCZ was a fixed dose of 400 mg initially, optimized to weight-based dosing of 8mg/kg later as new evidence got available. Patients were assessed clinically after the first dose. Also, inflammatory markers (CRP, ferritin) were repeated after 24-36 hours of the first dose. In the initial phase, IL-6 was checked post-TCZ in some of the patients to build knowledge about the levels of IL-6 after TCZ administration. One additional dose of TCZ was administered after 36-48 hours to those who had no improvement in terms of hypoxemia and inflammatory markers after the first dose. All the patients received low molecular weight or unfractionated heparin.

### Inclusion and exclusion criteria

Patients with a diagnosis of COVID-19 {based on a positive reverse-transcriptase polymerase chain reaction (PCR) test for SARS-CoV-2 virus; or presence of clinical signs and symptoms along with chest computed tomography (CT) result suggestive of COVID-19/ COVID-19 CT Classification (CO-RADS) 4 or 5}, who received TCZ during the study period were included in the study. Predefined exclusion criteria were active bacterial or fungal infection, ANC<500/mm^3^, PC<50000/ mm^3^, or SGPT/SGOT >5 times UNL.

### Data collection

Demographic details including age, sex, comorbidities, signs and symptoms; results of baseline and serial laboratory parameters, radiological investigations, ward, and intensive care unit (ICU) progress notes; and treatment and outcome details were obtained from the medical records files of the patients and hospital’s intranet. Status of respiratory support on all the days during hospitalization was recorded. Respiratory support was defined as per the following categories-1. No support, 2. Oxygen support with nasal cannula (O2 via NC), 3. Oxygen support with mask (O2 via mask), 4. Oxygen support with non-rebreathing mask (O2 via NRBM), 5. HFNC, 6. BiPAP 7. MV. Categories 2, 3, and 4 were considered low flow oxygen support. The details were recorded in hard copy as well as in an electronic database.

### Outcomes

The primary outcome was the need for invasive mechanical ventilation or death. Secondary outcomes were decrease in oxygen requirement as well as decrease in CRP, ferritin, D-dimer, and IL-6 after TCZ administration; secondary bacterial and fungal infections, and renal or hepatic dysfunction.

### Statistical analyses

Descriptive statistics were used for describing demographic and patient characteristics and were summarized as rates or percent for categorical variables, while median and 95% CI for median was used for the continuous variables. The change in continuous variables across the time point was assessed using the paired non⍰parametric Wilcoxon Signed⍰Rank Test. Kaplan Meier survival analysis was used for parameters such as age group, gender, and progression to mechanical ventilation by considering death as an event and log rank test was used for testing significance. Data were analyzed using IBM SPSS version 19.

## RESULTS

1081 patients with a diagnosis of COVID-19 were admitted between May 5 and July 31, 2020 in our hospital. 793 patients had mild disease. 545 patients received steroids, and 125 patients received TCZ along with steroids for treatment. We did not have any control group as TCZ was available in our hospital and was a part of the treatment protocol since we started treating COVID-19 patients. Some non-hypoxic patients received steroids for persistent fever with increasing inflammatory markers in the second week of illness. Data for 1 patient who received TCZ was not available and excluded in certain analyses. 10 patients left the hospital prior to their final outcome because of financial constraints or unavailability of bed; however, they were telephonically followed up to record their final outcome (7 patients recorded as discharged and 3 as died). Of the 125 patients who were treated with TCZ, 94 (75.2%) patients survived while 31(24.8%) patients died. The median duration between the onset of symptoms and receipt of TCZ was 10 days (95% CI 9.06 – 10). 100 patients received single dose of TCZ while 24 received 2 doses (1 patient data was lost). The interval between the first and second doses was 2 days (95% CI 2 - 3.3).

Table 1 shows the demographic characteristics of the patients. 80 % were male (n=100) and 20% were female (n=25). The median age amongst males and females was 55 years (95% CI 52.7 – 58.3) and 63 years (95 % CI 55.1 – 69.9), respectively. The youngest female was a 29 years old pregnant female patient, whereas the youngest male was 28 years. No other female patient was ≤ 40 years of age as compared to 11 male patients who were ≤ 40 years.

**Table 1:** Demographic characteristics of 125 patients who received TCZ.

| **Characteristic** | **Values** |
| --- | --- |
| **Median age, years, no. (95%CI)** | 56 (54-60) |
| **Sex, no. (%)** |  |
| Male | 100 (80) |
| Female | 25 (20) |
| **Comorbidities, no. (%)** |  |
| Hypertension | 64 (51.2) |
| Diabetes Mellitus | 54 (43.2) |
| Obesity | 32 (25.6) |
| Chronic cardiac disease | 17 (13.6) |
| Chronic pulmonary disease | 4 (3.2) |
| Chronic liver disease | 2 (1.6) |
| Chronic kidney disease | 4 (3.2) |
| Hypertension & diabetes mellitus | 17 (13.6) |
| Hypertension & obesity | 6 (4.8) |
| Diabetes Mellitus & obesity | 2 (1.6) |
| Hypertension, diabetes mellitus & obesity | 13 (10.4) |
| Hypertension, diabetes mellitus & chronic cardiac disease | 10 (8) |
| **Clinical features, no. (%)** |  |
| Fever | 120 (96) |
| Cough | 80 (64) |
| Dyspnea | 61 (48.8) |
| Bodyache | 29 (23.2) |
| Fatigue/weakness | 29 (23.2) |
| Sore throat/throat pain | 17 (13.6) |
| Headache | 11 (8.8) |
| Diarrhea | 9 (7.2) |
| Nausea/vomiting | 5 (4) |
| Dysguesia | 4 (3.2) |
| Abdominal pain | 4 (3.2) |
| Anosmia | 2 (1.6) |
| **Median hospitalization time for death, days, no. (95% CI)** | 13 (9.6 - 16) |
| **Median hospitalization time for discharge, days, no. (95% CI)** | 12 (10 - 13) |

The most common presenting symptom was fever (96%), followed by cough (64%) and dyspnoea (48.8%). The median duration of the first symptom, fever, and dyspnea prior to admission was 6 days (95% CI 5 - 6), 6 days (95% CI 5 - 7) and 3 days (95% CI 3 - 4) respectively.

78.4% patients had one or the other comorbidities (n=98), the commonest being hypertension, diabetes mellitus, and obesity. 12% of females (3/25) vs. 24% (24/100) males did not have any comorbidity. The median age of males without comorbidity was 51.5 years as compared to 57.5 years with comorbidity. The median age of females without comorbidity and with comorbidity did not have much variation. Some of the less common comorbidities were myasthenia gravis, epilepsy, hypothyroidism, chronic sinusitis, psoriasis, and multiple myeloma. Mortality rate in patients with hypertension was 31.3%, diabetes was 27.8%, and obesity was 25%. 50% patients died who had hypertension along with obesity. Only 2 (out of 27) patients without any comorbidities died. Mortality in patients with 2 comorbidities (41.7%) was twice the mortality in patients with only 1 comorbidity (20.9%).

Table 2 shows the level of respiratory support in 124 patients on the day of admission and the highest level of respiratory support during the hospital stay. 6 patients did not require oxygen support during the entire hospital stay; 3 patients amongst them had no hypoxia, and TCZ was administered for persistent high-grade fever in the second week of illness with increasing CRP (pre-TCZ absolute values of CRP were 61, 60 and 140 mg/L respectively) and/or high IL-6. The other 3 patients had a transient decrease in SpO2 on room air to 91-93% with high inflammatory markers and fever.

**Table 2:** Respiratory support on day of admission, day of TCZ and highest level of respiratory support during hospital stay (n=124)

| **Respiratory support** | **On day of admission**<br>no. (%) | **On day of TCZ**<br>no. (%) | **Highest level of Respiratory support any time during hospital stay**<br>no. (%) |
| --- | --- | --- | --- |
| Room air | 69 (55.6) | 12 (9.7) | 6 (4.8) |
| O2 via NC | 15 (12.1) | 43 (34.7) | 34 (27.4) |
| O2 via Mask | 9 (7.3) | 10 (8.1) | 7 (5.7) |
| O2 via NRBM | 16 (12.9) | 21 (16.9) | 13 (10.5) |
| HFNC | 3 (2.4) | 10 (8.1) | 13 (10.5) |
| BiPAP | 10 (8.1) | 20 (16.1) | 20 (16.1) |
| Mechanical Ventilation | 2 (1.6) | 8 (6.4) | 31 (25) |

Abbreviations: NC, nasal cannula; NRBM, non-rebreathing mask; HFNC, high flow nasal cannula; BiPAP, Bilevel positive airway pressure

Respiratory support in relation to the administration of steroids and TCZ, until the final outcome of the patient (discharge/death) was assessed. Number of patients on room air reduced from 70 on day of admission to 58 on the day steroids were started and further decreased to 12 on the day of TCZ. Though there was a trend towards an increase in the respiratory support up to 2 days after receipt of TCZ, on day 5 after TCZ, 62.1% patients had significantly improved (38.7% were on room air or discharged while 23.4% were on low flow oxygen). 65.3% patients were discharged by day 14. The longest hospital stay was 69 days (Table 3).

**Table 3:** Serial progression of respiratory support from day of admission till final outcome.

| *Respiratory support* | **Day of admission** | **Day of steroid** | **Day of TCZ** | **1 day after TCZ** | **2 days after TCZ** | **5 days after TCZ** | **7 days after TCZ** | **10 days after TCZ** | **14 days after TCZ** | **>14 days after TCZ** |
| --- | --- | --- | --- | --- | --- | --- | --- | --- | --- | --- |
| *Room air* | 70 | 58 | 12 | 10 | 11 | 35 | 30 | 9 | 4 | 3 |
| *O2 via NC* | 10 | 8 | 10 | 6 | 3 | 2 | 2 | 0 | 0 | 0 |
| *O2 via Mask* | 14 | 23 | 43 | 39 | 44 | 22 | 11 | 7 | 6 | 3 |
| *O2 via NRB* | 15 | 16 | 21 | 19 | 11 | 5 | 2 | 1 | 0 | 0 |
| *HFNC* | 3 | 3 | 10 | 16 | 20 | 10 | 7 | 7 | 0 | 0 |
| *BiPAP* | 10 | 11 | 20 | 22 | 16 | 15 | 8 | 6 | 1 | 1 |
| *Mechanical Ventilation* | 2 | 5 | 8 | 11 | 17 | 17 | 19 | 11 | 8 | 7 |
| ***Grand Total*** | **124** | **124** | **124** | **123** | **122** | **106** | **79** | **41** | **19** | **14** |
| ***Outcome*** |  |  |  |  |  |  |  |  |  |  |
| *Discharge* | 0 | 0 | 0 | 1 | 2 | 13 | 36 | 65 | 81 | 93 |
| *Death* | 0 | 0 | 0 | 0 | 0 | 5 | 9 | 18 | 24 | 31 |
| ***Total of outcome*** | **0** | **0** | **0** | **1** | **2** | **18** | **45** | **83** | **105** | **124** |

Numbers in the table represent the number of patients
Abbreviations: NC, nasal cannula; NRB, non-rebreathing mask; HFNC, high flow nasal cannula; BiPAP, Bilevel positive airway pressure

All patients who received TCZ on room air survived. Patients receiving TCZ on low flow oxygen had a mortality of 16.2%, and those on HFNC and BiPAP had a mortality of about 50%. Highest mortality was seen in patients on MV on the day of TCZ treatment (62.5%). Majority (71%) of patients died after day 7 of TCZ administration.

Table 4 and Fig 1 show the cumulative survival probability of patients as per age group, gender, and progression to MV. Fig 1 (A and B) show survival according to the age group (≤ 60 years and > 60 years) and gender, respectively; the difference in probability of the event (death) was statistically not significant (log rank test with p = 0.31and p = 0.28 respectively). Fig 1 (C) shows the survival related to progression to MV (required and not required); the difference in probability of the event (death) was statistically significant (log rank test with p < 0.0001).

**Table 4:**
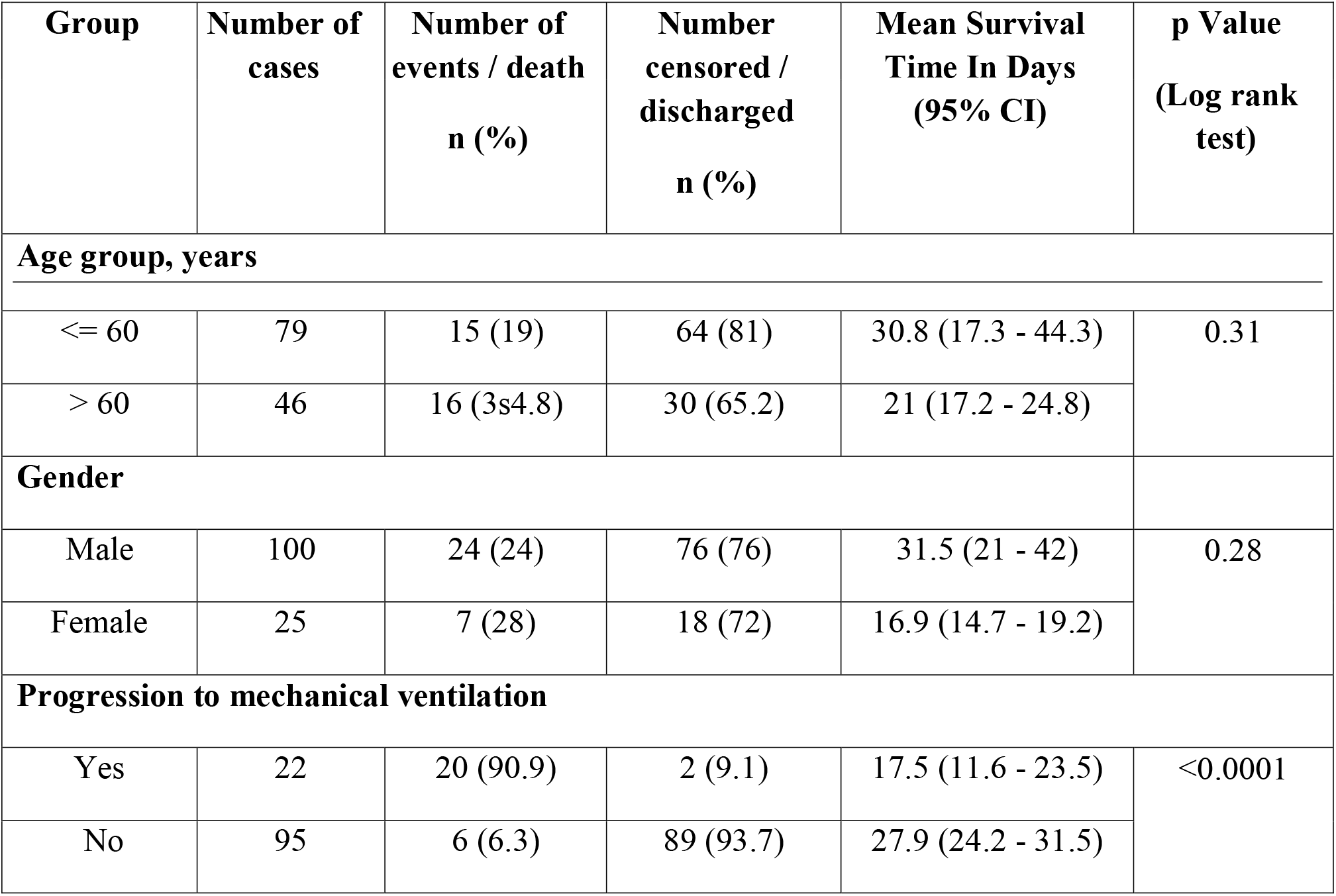
Survival analysis for age group, gender and progression to mechanical ventilation.

**Figure 1:**
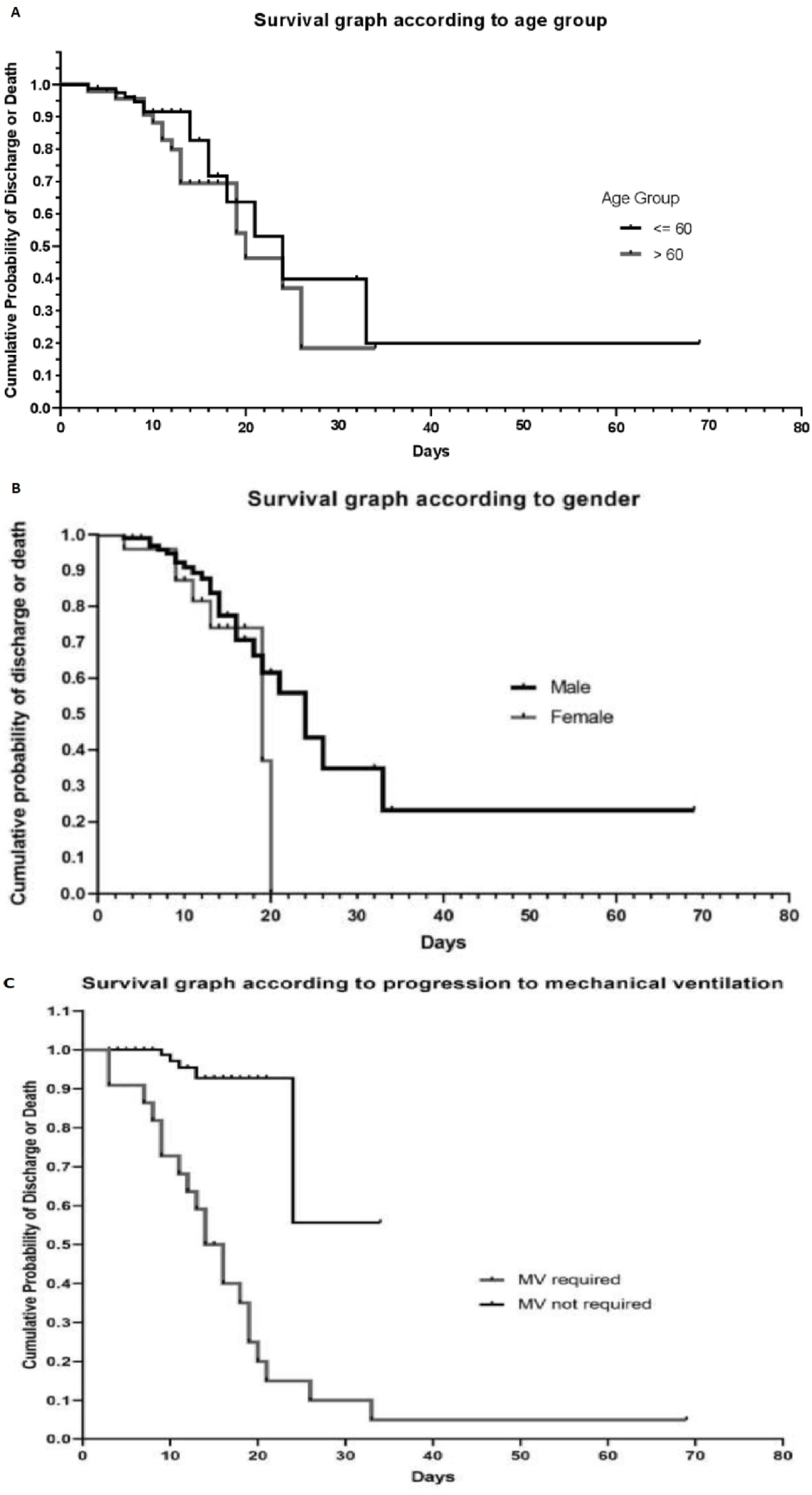
Cumulative probability of discharge or death according to age group (A), gender (B), and progression to MV for the event death amongst patients who were not on MV on day of TCZ (C).

On the day of admission, the median absolute lymphocyte count (ALC) was 1022 cells/cumm (95% CI 949.5 - 1087.9), neutrophil to lymphocyte (NL) ratio was 6.3 (95% CI 4.7 - 7.7), and serum lactate dehydrogenase (LDH) was 336.8 U/L (95% CI 260.5 - 382.8). Remaining laboratory parameters were serially assessed from the day of TCZ before the administration of the drug, till 7 days. Median CRP on the day of TCZ was 60.4 mg/L (95% CI 45.4 - 95.1), which decreased progressively to normal by day 5 after TCZ (Table 5). Table 6 shows the statistical association of various laboratory parameters after TCZ. There was a statistically significant decline in CRP on day 2, 5, and 7 as compared to baseline, and on day 5 and 7 compared to day 2.Maximum decline in CRP was attained by day 5. D-dimer and IL-6 increased after TCZ; the values of ferritin did not change significantly. No significant difference was found in the values of liver transaminases and creatinine.

**Table 5:** Serial laboratory parameters following TCZ administration.

| Lab parameter | On day of TCZ |  |  | 2 days after TCZ |  |  | 5 days after TCZ |  |  | 7 days after TCZ |  |  |
| --- | --- | --- | --- | --- | --- | --- | --- | --- | --- | --- | --- | --- |
|  | n | Median | 95% CI | n | Median | 95% CI | n | Median | 95% CI | N | Median | 95% CI |
| ALC (cells/cu.mm) | 116 | 821.9 | 778.5 - 937.3 | 110 | 903.2 | 781.2 - 1005.7 | 87 | 1294.2 | 1210 - 1481.1 | 65 | 1235.0 | 1020.4 - 1510.5 |
| N/L ratio | 116 | 11.4 | 10.8 - 12.9 | 110 | 9.5 | 8.3 - 10.8 | 87 | 6.4 | 4.5 - 8.2 | 65 | 7.7 | 6.2 - 9 |
| Creatinine (mg/dl) | 76 | 0.9 | 0.9 - 1.1 | 76 | 0.9 | 0.8 - 1 | 56 | 0.9 | 0.8 - 1 | 47 | 0.9 | 0.8 - 1.1 |
| ALT (U/L) | 68 | 35.3 | 27.5 - 47.1 | 63 | 40.1 | 32.9 - 62.3 | 49 | 50.9 | 40.6 - 70 | 36 | 56.1 | 39.1 - 74.2 |
| AST (U/L) | 26 | 39.6 | 32.9 - 54.5 | 15 | 43.3 | 31.7 - 106.6 | 7 | 50.7 | 31.9 - 486.5 | 8 | 57.3 | 27.2 - 386.3 |
| CRP (mg/L) | 108 | 60.4 | 45.4 - 95.1 | 99 | 20.6 | 16.4 - 29.6 | 82 | 4.1 | 3 - 6.3 | 54 | 2.8 | 1.963 - 3.6 |
| IL-6 (pg/ml) | 68 | 105.7 | 81.8 - 142.7 | 25 | 276.8 | 103.3 - 409 | 9 | 1000.0 | 215.4 - 1378.4 | 4 | 492.5 | - |
| Ferritin (ng/ml) | 77 | 562.3 | 475 - 718.3 | 57 | 646.0 | 482.5 - 921.8 | 44 | 591.6 | 499.1 - 903.5 | 33 | 580.8 | 425 - 801.3 |
| D-dimer (ng/ml) | 95 | 660.0 | 596.9 - 804.3 | 74 | 705.0 | 620 - 972 | 54 | 992.0 | 617.6 - 1363.1 | 38 | 1076.2 | 678.5 - 2099.7 |

Blank cells for 95% CI of IL-6 is due to very few observations on that day
SI conversion factors: To convert ALC to $\times 10^9/L$ , multiply values by 0.001
To convert creatinine to $\mu\text{mol/L}$ , multiply values by 88.4
To convert ALT and AST to $\mu\text{kat/L}$ , multiply values by 0.0167
To convert CRP to $\text{mg/L}$ , multiply values by 10
To convert ferritin to $\mu\text{g/L}$ , multiply values by 1
To convert D-dimer to $\text{nmol/L}$ , multiply values by 5.476
Abbreviations: ALC, absolute lymphocyte count; N/L ratio, neutrophil lymphocyte ratio; ALT, alanine aminotransferase; AST, aspartate aminotransferase; CRP, C-reactive protein; IL-6, interleukin 6

**Table 6:** Association of laboratory parameters on various days following TCZ administration.

| Lab parameter | n | Baseline<br>vs day 2<br>(p) | n | Baseline<br>vs day 5<br>(p) | n | Baseline<br>vs day 7<br>(p) | n | Day 2 vs<br>day 5 (p) | n | Day 2 vs<br>day 7 (p) | n | Day 5<br>vs day<br>7 (p) |
| --- | --- | --- | --- | --- | --- | --- | --- | --- | --- | --- | --- | --- |
| ALC (cells/ cu.mm) | 103 | 0.6342 | 80 | <<br>0.0001 | 60 | 0.0001 | 81 | < 0.0001 | 62 | < 0.0001 | 64 | 0.9116 |
| N/L ratio | 103 | 0.2 | 80 | <<br>0.0001 | 60 | 0.017 | 81 | < 0.0001 | 62 | 0.0023 | 64 | 0.14 |
| Creatinine (mg/dl) | 60 | 0.27 | 40 | 0.68 | 34 | 0.8 | 41 | 0.98 | 34 | 0.77 | 36 | 0.24 |
| ALT (U/L) | 46 | 0.04 | 30 | 0.006 | 22 | 0.16 | 33 | 0.83 | 27 | 0.31 | 27 | 0.006 |
| AST (U/L) | 10 | 0.93 | 6 | 1 | 5 | 0.125 | 6 | 1 | 6 | 0.62 | - | - |
| CRP (mg/L) | 91 | <<br>0.0001 | 71 | <<br>0.0001 | 48 | <<br>0.0001 | 69 | < 0.0001 | 45 | < 0.0001 | 51 | 0.109 |
| IL-6 (pg/ml) | 10 | 0.109 | 8 | 0.003 | - | - | 6 | 0.25 | - | - | - | - |
| Ferritin (ng/ml) | 49 | 0.491 | 32 | 0.55 | 25 | 0.0425 | 33 | 0.039 | 25 | 0.0089 | 27 | 0.92 |
| D-dimer (ng/ml) | 61 | 0.0706 | 24 | 0.0047 | 28 | 0.0001 | 26 | 0.0727 | 31 | 0.001 | 29 | 0.605 |

Blank cells for the statistical association of AST and IL-6 are due to very few observations on that day
Non-parametric Wilcoxon Signed Rank test is used for statistical analysis

Blood and lower respiratory cultures were ordered in 52 and 11 patients respectively. 12/52 blood cultures and 9/11 respiratory cultures were positive (8 out of the 9 patients with positive respiratory cultures had blood cultures positive too). Organisms which were isolated were *acinetobacter species, klebsiella pneumoniae, enterococcus species, stenotrophomonas maltophilia, pseudomonas aeruginosa and candida albicans/non-albicans*. All these patients had severe disease and were admitted in ICU. 11 out of 12 patients with positive blood cultures died. The rate of secondary infection in our study based on the positive blood and respiratory culture data was 10.4%.

## DISCUSSION

C3G study highlights the role of TCZ in COVID-19 with CRS. In this study, receipt of TCZ was associated with low occurrence of the primary outcomes i.e. death and need for mechanical ventilation in patients who had features of host-inflammatory response related injury. It is crucial to identify CRS at its onset. Corticosteroids are being used as the first-line anti-inflammatory agents in COVID-19 after the results of the RECOVERY trial.^10^ However, there are patients with suboptimal response to steroids. In our study, in patients who did not show adequate and timely response to steroids, administering TCZ prior to progression of cytokine storm necessitating HFNC, BiPAP or MV; was associated with low mortality.

The factors to be considered before administering TCZ are duration of the illness, response to steroids, the inflammatory markers, and the presence of bacterial or fungal infection.

Inflammatory markers like CRP and IL-6 have been found to be associated with disease severity and prognosis.^11-14^ The statistically significant decline in CRP post-TCZ reflects its role in assessing the response to the drug. The maximum decline in CRP by day 5 found in our study, is consistent with the results of the CORIMUNO TOCI 1 trial.^7^ Various studies have described a correlation between concentrations of ferritin, D-dimer, and LDH on admission with the severity of COVID-19.^15^ In this study, they did not show any significant reduction after TCZ administration acutely and hence may not help in judging the response to treatment. Similarly, IL-6 levels increased after the administration of TCZ due to blockade of receptors.

The incidence of secondary infections has varied in different studies.^16-18^ In our study, no patient admitted in the ward had positive blood culture, which indicates that sepsis after TCZ is more likely to occur in very sick patients with prolonged hospitalization in the ICU and maybe a contributor to mortality. No other major adverse events were noticed in our study.

The efficacy of TCZ has been discussed in various studies, though the timing of administration has varied.^16,19^ A recent meta-analysis found 12% lower mortality in patients treated with TCZ compared to those who were not.^20^ Our study emphasizes the importance of timing of TCZ administration and its association with the final outcome. The mortality in our study was least in patients with transient hypoxia and those on low flow oxygen. The outcome was poorest in patients who were on MV on the day of TCZ administration. Survival benefit was observed in patients who received TCZ while in respiratory support categories 1 – 4, by preventing progression to MV. The broad selection criteria and study design factors in the COVACTA trial seemed to have limited the utilization of TCZ strategically in clinically different subpopulations of patients, thereby necessitating cautious interpretation of the results.^21^

In the CORIMUNO TOCI 1 trial, even if the mortality on day 28 was not reduced, the decreased need for MV at day 14 would translate into lesser acute and long term complications associated with prolonged ICU care. During the ongoing pandemic, when there is a dearth of ICU beds in resource-limited settings, TCZ could shorten the hospital stay and be cost-effective. We support the strategy of combined use of steroids and TCZ in moderate to severe COVID-19, targeting the different arms of the inflammatory cascade.^19, 22^

### Limitations

Our study has many limitations. First, observational studies cannot draw causal inferences because of several confounders, especially absence of a control arm. However, in our viewpoint, a control group would not be rational, considering the role of TCZ in the treatment of CRS, which is a life-threatening condition. Second, misclassifications of data are possible because this was a retrospective cohort study, and data was manually extracted. Third, treatment with TCZ was done at the discretion of treating clinicians, despite a predefined protocol. Fourth, the detailed analysis of timing and doses of steroids is not done in this study, which may be an important factor towards the outcome.

Further studies are needed to define the subgroup of patients who would benefit most with the combination of steroids and TCZ, including the optimal timing of administration of both the drugs.

## CONCLUSION

Use of TCZ in Covid-19 patients with features of CRS is associated with decreased need for invasive ventilatory support and reduced occurrence of death as it may prevent the progression of the disease; without any major adverse events. The optimal timing of administration of TCZ, in the early CRS or hypoxemic phase, where there is clinical worsening despite administration of steroids, may be one of the key factors towards the final outcome.

## Supporting information

Strobe Checklist for Cohort study

## Data Availability

No URL for external data sets or online supplementary material pertain to this manuscript.

## Notes

### Author Contributions

Surabhi Madan and Manish Rana conceived and designed the study and wrote the manuscript. Amit Patel approved the final manuscript. Manish Rana and Nirav Bapat performed the analysis. Surabhi Madan, Amit Patel, Nitesh Shah, Vipul Thakkar, Bhagyesh Shah, Pradip Dabhi, Minesh Patel, Hardik Shah, Rashmi Chovatiya, Maulik Soni, and Rohan Gajjar collected and contributed data. Rohan Gajjar performed and supervised data compilation in electronic database. All authors read and approved the final manuscript.

## ACKNOWLEDGMENT

The authors thank Dr Parloop Bhatt, Dr Riddhi Parikh, Dr Disha Patel, Kartikae Sharan, Shayon Ghosh, Maitri Shah, Vishnu Venugopal, Sangeetha Arunachalam, Aayushi Singh; Department of Clinical Research, Care Institute of Medical Sciences, Ahmedabad, India, for their assistance in data aggregation and compilation in electronic database.

